# Population Estimates of People with Spina Bifida in the United States in 2020

**DOI:** 10.1101/2022.02.03.22268639

**Authors:** Virginia G. Briggs

## Abstract

**Introduction:** Spina bifida is a birth defect that results in deficits of neurological function. Individuals diagnosed with spina bifida often require a lifetime of medical care to manage this condition. Currently, the number of people living with spina bifida in the United States is unclear. The purpose of this study is to provide estimates of the magnitude of this condition, and its distribution by gender and age.

**Methods:** Total births affected by spina bifida were calculated using rates from the Birth Defects Monitoring Program (BDMP) and state-based birth defects tracking systems supported by the Centers for Disease Control (CDC), over an 80-year period (1940-2020). Spina bifida mortality rates were determined using death certificate data available through the Centers for Disease Control and Prevention, National Center for Health Statistics. Life tables were created for each year of birth between 1940 and 2020 to estimate the total number of people with spina bifida alive in 2020 in the United States.

**Results:** In 2020, the estimated number of people in the U.S. living with spina bifida (0-80 years of age) was 124,150 (67,662 female; 56,488 male). The majority were adults aged 30 to 80 years (66.6%), compared to children and young adults (33.4%).

**Conclusion:** Understanding the approximate size and distribution by age and gender may assist health care providers in planning services for this changing population.

## 1. INTRODUCTION

Spina bifida (SB) is a birth defect characterized by the failure of the spine to close during fetal development, often resulting in damage to the spinal cord and surrounding nerves (1). Rates of SB in the United States (U.S.) began declining in 1996, when fortification of grains with folic acid was optional and continued after 1998 when fortification became mandatory (2).

Spina bifida is often accompanied by hydrocephalus as well as deficits in mobility, balance, bladder and bowel function. People with spina bifida experience health complications throughout their lifetime, including shunt malfunction, tethered cord syndrome, paralysis, bowel and bladder incontinence, skin ulcers and other problems requiring health care management and/or treatment (3). Mortality in children with spina bifida has been reported to be as much as ten times that of children without birth defects (4). Mortality in adults with spina bifida is most often associated with infection, respiratory failure, renal failure and shunt malfunction (5). However, because of advances in medical management of some of these associated health conditions, survival has improved dramatically (6). As a result of these changes in survival, the population size and age distribution of the spina bifida population has likely changed over time.

To better understand and plan for the care of these individuals now and in the future, health care systems in the United States would benefit from learning more about this population. Currently, the number of individuals living with spina bifida in the United States is unclear. Previous estimates have reported that over 160,000 people with spina bifida live in the United States (7, 8). In 2001, Larson, et al. used data from the National Health Interview Survey (NHIS) collected in 1994 and 1995 to produce a weighted estimate of the spina bifida population in the U.S. (8). Two considerations for the accuracy of this estimate are that the NHIS did not use Simple Random Sampling (SRS), a method best for estimating numbers of people with a specific condition (9), and the estimate used data collected 25 years prior to this study, providing outdated information about the SB population size.

Reported rates of spina bifida births in the literature have fluctuated since the 1990’s, primarily due to the increase of folic acid in the U.S population diet, but also due to the varying methods used to document births affected by spina bifida (10). Previous rates have used birth certificate information to calculate the rate of spina bifida affected births (11). Birth certificates have been shown to be an unreliable source for the identification of spina bifida affected births compared to other surveillance systems (12). Another method of identifying spina bifida cases has been the inclusion of still births and terminated pregnancies in addition to live births, which would overestimate the number of people living with spina bifida if they were not excluded from the estimates. The purpose of this study is to estimate the current size of the spina bifida population in the United States, by identifying live births affected by spina bifida and adjusting for mortality over an 80-year period, and to determine the gender and age distribution of these individuals.

## 2. METHODS

### 2.1. Total Births in the United States

Estimates for total births for all included years were attained from the Vital Statistics of the United States Natality Reports produced by the National Center for Health Statistics (13, 14). The estimates consisted of all documented live births in all states on an annual basis between, 1940 and 2020. Eighty years was chosen as the cutoff since life expectancy in the U.S. is 78.7 years (15).

### 2.2. Spina Bifida Birth Rates

Rates used to calculate the number of spina bifida affected births each year for this study were taken from surveillance systems that did not utilize birth certificates for their estimates, were from nationally representative samples and reported information on live births. Rates reported in the Birth Defects Monitoring Program (BDMP) were used for the years 1970-1994 (16,17,18,19). From 1970 to 1982, rates reported in the BDMP combined still births and live births. To extract live birth estimates for this study in these years, still births were deducted from the BDMP rates using still birth risk estimates associated with spina bifida reported by Heinke, et al. (20), of 32/1,000. As no birth rates for spina bifida were available for the years 1940-1969, the mean rate for the first two years of data available from the BDMP (1970 and 1971) was applied to all years before 1970. From 1994 to 2014, rates were used that were based on state surveillance systems (that excluded stillbirths and terminated pregnancies) and reported by Williams, et al. (21,22) and Mai (23). As no birth rates for 2015-2020 were available, the same rates were applied that Mai, et al. reported for years 2010-2014 (3.3/10,000). A description of these sources, by year, is shown in Appendix 1.

### 2.3. Spina Bifida Mortality

Mortality for people with spina bifida was calculated using the life table approach. Using death certificate data collected by the National Center for Health Statistics (NCHS) and available through the Centers for Disease Control’s Wide-ranging Online Data for Epidemiologic Research system (WONDER), life tables were constructed consisting of each year of birth in the study period (24,25,26). Causes of death were documented differently in three separate data collection periods. From 1968 to 1978, only underlying causes of death were reported and were classified using the Eighth Revision of the International Classification of Disease Adapted for Use in the United States (ICDA-8; Codes 740, 740.0 and 740.9). From 1979-1998, underlying causes of death were also reported and grouped by age, but were classified using the International Classification of Disease Ninth Division (ICD-9; Codes 740, 740.0 and 740.9). For both these periods, deaths were reported by age group (<1, 1-4, 5-9, 10-14, 15-19, 20-24, 25-34, 35-44, 45-54, 55-64, 65-74 and 75-84). To provide the closest estimate of number of deaths for each single-year age, the number of deaths for each age group was divided evenly between each year of age included. For example, if 100 deaths were reported for the age 5-9 group in one year, then each of the 5 years of age would be assigned one-fifth the number of deaths (i.e. 20 children died at age 5 years, etc.). Finally, from 1999-2020, multiple causes of death were reported providing the ability to extract deaths where any mention of spina bifida was given. Deaths were available for single-year ages rather than only age groups, and were classified using the International Classification of Disease Tenth Division (ICD-10; Codes Q05, Q05.0-Q05.9). Cause of death information was not available for the years 1940 through 1967. Spina bifida-related deaths occurring within this timeframe were estimated using the proportion of deaths (by age) associated with spina bifida in individuals born in the first two years of available data (1968-1969). This method was also used by Presson, et al. in a similar study in 2010 examining the population size of individuals with Down Syndrome in the United States (27). A description of these sources, by year, is shown in Appendix 2.

### 2.4. Spina Bifida Population Estimates by Age and Gender

Calculations of the number of people living with spina bifida by gender were made using the ratio of the female to male prevalence rate (1.2) in people with spina bifida, reported by Lary, et al. (19). This ratio was determined in both state surveillance systems and the Center for Disease Control’s Birth Defects Monitoring Program (BDMP). The age distribution of people with spina bifida alive in 2020 was determined by using the life tables constructed for each year of birth.

## 3. RESULTS

A total of 161,912 live births affected by spina bifida were estimated to have occurred in the United States between 1940 and 2020, of which 124,150 survived to the end of 2020 (Table 1). The distribution by gender included 67,662 (54.5%) females and 56,488 (45.5%) males. Children and young adults accounted for 33.4% (n=41,525) of the SB population (age 0 to 29 years) while 66.6% (n=82,625) were adults, age 30 to 80 years.

**Table 1:**
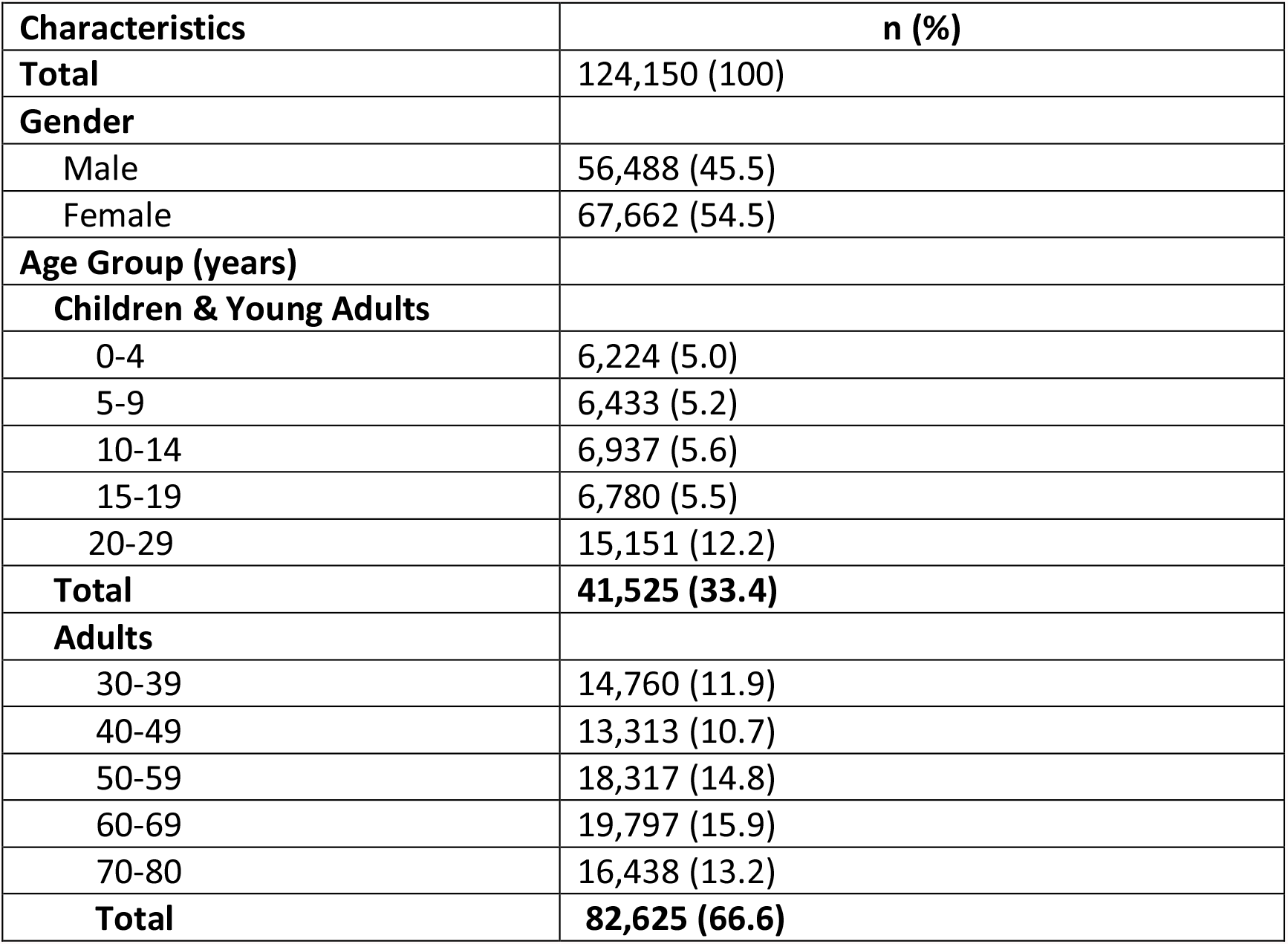
Population Estimates of People (0-80 years) with Spina Bifida in the U.S. in 2020.

## 4. DISCUSSION

The estimates reported in this study have some limitations, as several assumptions were made in estimating both the number of births and number of deaths associated with spina bifida. In some years during the study period, data were either missing or limited for SB affected births and/or deaths. This was particularly evident in the earlier years of the study period, from 1940-1969. The first problem was the limited documentation of SB affected births, especially in a nationally representative sample. It is possible the SB birth rates in this 29-year period were very different from the early 1970’s, especially for years around World War II when nutrition may have been poor. Thus, applying one rate for all years before 1970 may not accurately represent the number of spina bifida affected births in that time period. In a meta-analysis by Atta, et al. (10), rates of SB births were examined worldwide, showing wide variation geographically as well as within the folic acid pre-fortification years. However, the SB birth rates used for the current study were chosen based on the methods used to calculate them, which included nationally representative study samples in the U.S. that included information on live births, and were identified by methods other than birth certificate information. For consistency in this study, applying SB birth rates that fit this criteria to the closest years with little or no documentation likely provided the closest estimates. The separation of stillbirths and live births in published rate calculations was also a problem in some years of surveillance. Though the stillbirth rate for SB reported by Heinke, et al. (20) was used to extract the number of live births in 1982 and earlier in this study, this rate was calculated using data from a later time period of 1997-2011. If stillbirth rates were higher in the earlier years, then the number of stillbirths would have been underestimated.

The second problem was the difference in documentation of mortality over time. First, since only the underlying cause of death was documented between 1968 and 1998, some death certificates may have listed another cause of death in people with spina bifida, which would underestimate the number of spina bifida-related deaths. Second, deaths were grouped by age, giving one single number of deaths for each age group. Dividing this single number evenly between each year of age may have affected the overall number of deaths. However, as mortality over a lifespan is difficult to assess in a medically compromised population, it was important to adjust for both age-related and spina bifida-related deaths using a life table approach. The third consideration concerning mortality estimates was the lack of cause-of-death information from 1940 to 1967. Many individuals were still alive in 2020 who were born within this timeframe, and it is vitally important to know more about them, but difficult to assess mortality without specific information that was not yet available. Though the lifespans of people born with spina bifida in these earlier years were assumed to have the same mortality rates as people born in 1968 and 1969 for the purposes of this study, improvements in health care over time could have also caused an under-estimate of deaths between 1940 and 1967.

Finally, since this study used only births and deaths of people with spina bifida in the United States, those who came to the U.S. after birth were not captured in the birth estimates. For example, the Hispanic/Latino population has been growing steadily in the U.S. in the last ten years (28). Given the high rates of spina bifida in this population (29), there are likely cases of spina bifida that arrived in the U.S. after birth, and not included in the estimate reported in this study. Similarly, those that died outside the U.S. would not have been captured in the mortality estimates used to calculate the current SB population size.

The estimates reported in this study are an update to previously reported SB population sizes in the U.S., and are a starting point for health care providers to plan for the care of individuals with spina bifida living in the United States today. Though age and gender distribution are important, other characteristics may also be useful for understanding where health care resources should be allocated for this unique population. Condition severity, comorbidities, social support systems, language barriers, mental health status and other characteristics may also assist health care providers in determining medical care and social services for individuals with spina bifida of all ages. As medical care improves and lifespan increases, the needs of this population will continue to change.

Future estimates may include a more comprehensive examination of spina bifida affected live births in years prior to folic acid fortification of grains. Additional studies may also include assessment of U.S. residents with spina bifida born outside the U.S. By refining the documentation methods used to identify spina bifida affected births in earlier years and associated rates of death, more accurate population estimates will allow health care providers to better prepare for their care.

## Data Availability

All data used for this study are available online at:
1)https://www.cdc.gov/nchs/data-visualization/natality-trends/index.htm
2)http://wonder.cdc.gov/cmf-icd8.html
3)http://wonder.cdc.gov/cmf-icd9.html
4)http://wonder.cdc.gov/mcd-icd10.html

http://wonder.cdc.gov/cmf-icd8.html

http://wonder.cdc.gov/cmf-icd9.html

http://wonder.cdc.gov/mcd-icd10.html

## Acknowledgements

No funding sources were used to support this research.

---

**Appendix 1:**
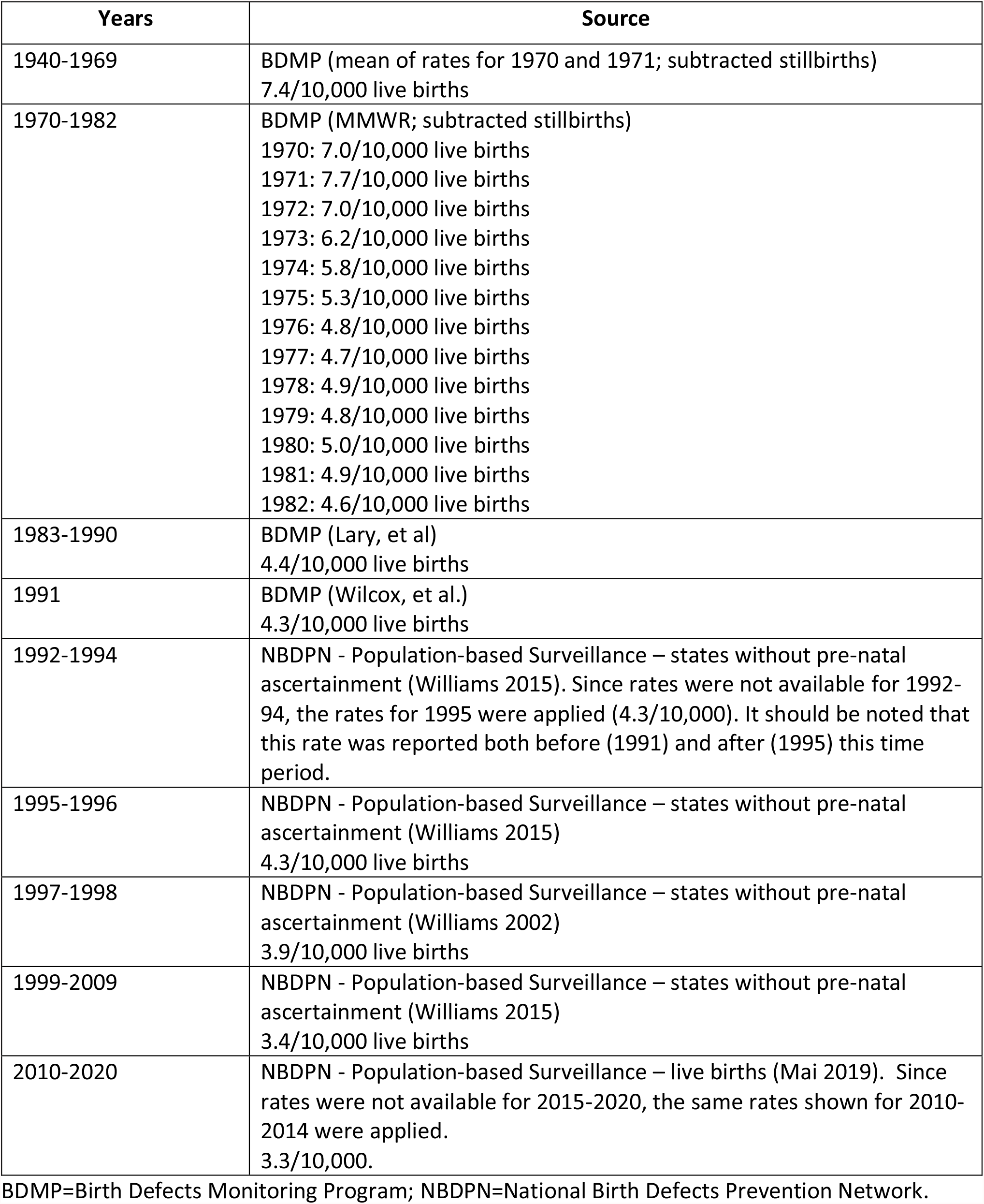
Source of spina bifida affected live birth rates used in estimates.

**Appendix 2:** Source of spina bifida affected mortality rates used in estimates.

| **Years** | **Source** |
| --- | --- |
| 1940-1967 | Used the proportion of deaths (by age) associated with spina bifida in individuals born in the first two years of available data (1968 and 1969). Based on ICDA-8 codes and on underlying cause of death only. |
| 1968-1978 | Based on ICDA-8 codes and on underlying cause of death only. Deaths were reported by age group. |
| 1979-1998 | Based on ICD-9 codes and on underlying cause of death only. Deaths were reported by age group. |
| 1999-2020 | Based on ICD-10 codes and on multiple causes of death. Deaths were reported by single age. |

